# Association Between Psychiatric Polygenic Scores, Healthcare Utilization and Chronic Disease Comorbidity Burden Among European Ancestry Individuals

**DOI:** 10.1101/2023.09.29.23296345

**Authors:** H. Lester Kirchner, Daniel Rocha, Richard K Linner, Drew Wilimitis, Colin G Walsh, Michael Ripperger, Hyunjoon Lee, Zhaowen Liu, Lea Davis, Yirui Hu, Christopher F Chabris, Jordan W Smoller

## Abstract

**Purpose:** To estimate the association of psychiatric polygenic scores (PGS) with healthcare utilization and comorbidity burden.

**Methods:** Observational cohort study (N=118,882) of biobank participants 15+ years of age from three diverse healthcare systems. PGS derived for major depressive disorder (MDD), bipolar disorder, and schizophrenia, were tested for association with healthcare utilization (measured as frequency of emergency department (ED), inpatient (IP), and outpatient (OP) visits) and comorbidity burden (defined using Elixhauser and Charlson Comorbidity Indices).

**Results:** Individual health records had a median EHR duration of 12 years. Those in the top decile of MDD PGS but without diagnosed MDD had significantly more ED visits (RR=1.22, 95% CI; 1.17, 1.29) compared to those in the bottom decile. Increases were also observed with IP and comorbidity burden. Among those diagnosed with depression and in the highest decile of the PGS, there was an increase in all utilization types (ED: RR=1.55, 95% CI 1.41, 1.72; OP: RR=1.16, 95% CI 1.09, 1.23; IP: RR=1.23, 95% CI 1.12, 1.36) post-diagnosis. A few statistically, however no clinically, significant results were observed with bipolar and schizophrenia PGS.

**Conclusions:** MDD PGS is associated with increased utilization and comorbidity burden, even in the absence of a depression diagnosis. Following a diagnosis of depression, the MDD PGS was associated with further increases in utilization. These findings suggest that MDD PGS may serve as a biomarker of health outcomes in real-world settings.

## INTRODUCTION

Mental health care is paramount in the management of chronic medical conditions that co-occur with serious mental illness (SMI) including major depressive disorder (MDD), bipolar disorder (BD), and schizophrenia (SCZ)^1^. For example, in a Canadian cohort of adults with at least one chronic medical condition (asthma, congestive heart failure, myocardial infarction, diabetes, epilepsy, hypertension, chronic pulmonary disease, and chronic kidney disease), those with comorbid MDD or SCZ were found to have an increased mean total 3-year cost of care compared to those without any co-morbid mental health disorder^2^. Mental health and substance abuse treatment costs among individuals with BD and MDD were two and three times, respectively, as high as general medical outpatients^3^. Several other studies have found increased costs and utilization among patients with co-morbid medical disease and depression^4-7^.

In parallel, mounting evidence indicates that genetic factors can increase the risk of both SMI and chronic medical conditions^8,9^. For example, prior work from the PsycheMERGE network, a multi-center network of electronic health record-(EHR-) linked biobanks, demonstrated that polygenic scores (PGS) for MDD also increase the risk of cardiovascular disease even in patients without diagnosed depression^10^. Moreover, depression is a modifiable risk factor for cardiovascular disease^11^. Other findings from PsycheMERGE have shown that a PGS for SCZ is associated with a range of non-psychiatric medical conditions, including hepatitis, neurologic disorders, and disorders of the urinary system, though not with cardiovascular or metabolic dysfunction^12^. This raises the hypothesis that polygenic scores for SMI may be associated with comorbidity burden or health care utilization, even in the absence of SMI. If so, this could have implications for selecting effective models of care that reduce comorbidity and health care costs. For example, the collaborative chronic care model (CCM) takes a team-based patient-centered approach and has demonstrated efficacy in reducing comorbidities for patients with SMI^13,14^. Recent genetic findings described above suggest that a CCM which integrates psychiatric and mental health care may be a viable *preventative* model of care for patients with high genetic risk of psychiatric illness, even in the absence of SMI.

A large body of research has established that psychiatric PGS capture significant risk of psychiatric disorder, largely in cohorts ascertained for research. However, the prospect of implementing polygenic scores as risk indices in real-world health settings has been understudied. In addition, little is known about whether PGS are associated with disease burden and care utilization. The observation that genomic risk for psychiatric disorders is correlated with a range of medical outcomes suggests that psychiatric PGS may be associated with multimorbidity, and consequent healthcare service use in absence of risk for a PGS’s cognate disorder.

In this study, we leverage the PsycheMERGE network to examine whether polygenic scores for major psychiatric disorders (MDD, BD, and SCZ) are associated with important clinical outcomes: healthcare utilization and comorbidity burden. We focused on the utilization outcomes related to inpatient, outpatient, and emergency department usage, and the number of comorbid conditions as indexed by two widely used measures: the Elixhauser Comorbidity Index (ECI)^15^ and the Charlson Comorbidity Index (CCI)^16^.

## MATERIALS AND METHODS

### Participating Sites

Participating study sites included two major academic medical centers and a large rural healthcare system in the United States; Mass General Brigham (MGB), Vanderbilt University Medical Center (VUMC), and Geisinger. Each site includes a hospital-based biobank; the MGB Biobank^17^, the VUMC biobank (BioVU)^18^, and the Geisinger MyCode Community Health Initiative (MyCode)^19^. At each of these sites, participants were recruited from the general healthcare system population without systematic recruitment for any specific disease or diagnosis. Biobank procedures and patient recruitment were approved at each health system’s institutional review board (IRB). Each biobank consent allows for EHR data linking and analysis of de-identified data. The IRB at the three sites approved the use of participant data in this analysis. For this study, only European ancestry participants aged 15 or older with at least three encounters over a period of at least five years in the EHR were included. All available historical EHR data were included at each site.

### Psychiatric Conditions

#### Major Depressive Disorder (MDD) and Schizophrenia (SCZ)

Cases of MDD and SCZ were identified by the presence of Phecodes for these conditions in their medical record. Phecodes were mapped from International Classification of Diseases (ICD)-9/-10 diagnosis codes using Phecode Map 1.2^20^. A case was defined as positive if the corresponding Phecodes were present on at least two outpatient (OP) visits at least 30 days apart, or at least one emergency department (ED) visit, or at least one inpatient (IP) visit, or present on the Problem List (PL), where available. The ICD9 and ICD10 codes used for MDD and SCZ are listed in Tables S1 and S2.

#### Bipolar Disorder (BD)

Cases of BD were defined using a rule-based phenotype algorithm developed at MGB as part of the International Cohort Consortium for Bipolar Disorder and referred to as “Bipolar Coded-Broad”^21^. This rule-based algorithm demonstrated high positive predictive value (0.80) against direct-interview clinician assessment based on the SCID-IV. To meet Bipolar Coded-Broad criteria, cases had at least two BD diagnostic (ICD) codes recorded at any time with a minimum of four weeks between each code and a history of two or more medications used to treat BD within one year of index BD diagnosis. To rule out patients with closely related disorders, the algorithm required the number of diagnostic codes for MDD, SCZ, Schizoaffective disorder, or Organic Affective Syndrome to total fewer than half the number of BD codes.

#### Polygenic Scores

To generate PGS, we used publicly available summary statistics from the Psychiatric Genomics Consortium’s (PGC) genome-wide association studies (GWAS) for MDD^22^, SCZ^23^, and BD^24^, respectively. VUMC used the latest available GWAS for BD by the Psychiatric Genomics Consortium (PGC)^25^ as these results were available when VUMC ran their analyses. To derive posterior GWAS coefficients adjusted for linkage disequilibrium (LD), we applied PRS-CS (September 10, 2020, software release), applying default settings^26^. For computational feasibility, PRS-CS is restricted to 1,120,697 quality-controlled HapMap 3 SNPs. PGS were computed for each participant by summing genotypes weighted by the posterior coefficients using PLINK1.9, build 6.17^27^. We provide details on the genotyping and PGS quality-control procedures in the **Supplement**.

### Outcomes

#### Healthcare utilization

The number of ED, IP, and OP visits were enumerated for each participant across the span of their EHR from the age of 15 to date of the data extraction. An ED visit was defined as only those that did not result in a hospital admission. OP visits included ambulatory in-person and telehealth encounters.

#### Comorbidity burden

Comorbidity burden was measured using the Elixhauser Comorbidity Index (ECI)^15^ and the Charlson Comorbidity Index (CCI)^16^ as indicators of how ill the participant was at the end of their EHR span. The ECI consists of 31 conditions and the CCI consists of 17 conditions (Table S3). A given condition was coded as positive following the same rules used to define MDD and SCZ. The ECI includes psychosis and depression as two of the conditions which may induce a correlation with the psychiatric PGS. Results were compared when including and excluding these two conditions and found to be similar. Therefore, results are shown for the full ECI.

#### Statistical Analysis

Descriptive statistics are presented as median and inter-quartile range for continuous variables, and frequency and percentages for categorial variables for each participating site. To estimate the effect of psychiatric PGS on healthcare utilization and comorbidity burden the negative binomial (NB) regression model was used. The NB distribution fit better than the Poisson as visually assessed by a rootogram^28^ and compared using a likelihood ratio test. To account for the varying lengths of EHR follow up time, an offset term, the natural logarithm of the EHR duration in years, was included in the models to allow for the standardized interpretation as the average number of visits per year. The models also controlled for participant age at the end of their EHR span, the first ten ancestry principal components derived from GWAS data, and genotyping batch. Analyses were performed separately at each site and results combined using an inverse-variance weighted meta-analysis. PGS were standardized at each site and treated as a continuous predictor as well as categorized into deciles. For the latter, the primary comparison was between the highest and lowest decile.

Participants meeting the definition of MDD, SCZ, or BD were censored on the day prior to the first instance of a corresponding diagnosis code. That is, the calculation of healthcare utilization and comorbidity burden only used EHR data up until the date of censoring while still requiring a minimum of 5 years in the health system. Thus, no EHR data from after a diagnosis of any of these psychiatric conditions were used in the primary analysis. For participants that did not receive a diagnosis of these conditions, all data in the EHR after the age of 15 were used to calculate the outcomes. We also conducted a subgroup analysis to estimate the association between MDD PGS and healthcare utilization after the diagnosis of depression. Lastly, a sensitivity analysis was conducted at one site (Geisinger) excluding participants meeting the definition of MDD, SCZ, or BD and the primary analyses were repeated. This was performed to remove any potentially induced associations between the PGS and the outcomes due to a delay in diagnosis where the patient may have incurred an increase in healthcare utilization associated with screening and assessment for a SMI. Results are presented as rate ratios (RR) and 95% confidence intervals (CIs). Analyses were performed using R v4.0.3^29^ with the countreg^30^ and MASS^31^ packages.

## RESULTS

A total of 111,991 biobank participants were included in the MDD PGS analysis, and 118,882 in the SCZ and BD PGS analyses. The majority were female (53%-58%) and median duration in the EHR was at least 12 years in all sites (Table 1). Additional summary information are presented in Table 1 and Table S4.

**Table 1:** Descriptive Statistics of Major Depressive Disorder Polygenic Score in the Depression Cohort.

| Variable | Site 1<br>(N=51,505) | Site 3<br>(N=24,826) | Site 2<br>(N=35,660) |
| --- | --- | --- | --- |
| Age, median (IQR)<sup>a</sup> | 58.94 (17.17)<sup>a</sup> | 61.77 (15.40) | 59.22 (16.59) |
| Female Sex, n (%)<sup>b</sup> | 30,072 (58.4%) | 9,804 (53.1%) | 20,686 (58.0%) |
| EHR Duration (years), median (IQR) | 14 (10, 19)<sup>b</sup> | 15 (9, 20) | 12 (8, 16) |
| Emergency Department Visits, median (IQR) | 1 (0, 3) | 0 (0, 2) | 0 (0, 1) |
| Inpatient Visits, median (IQR) | 0 (0, 1) | 5 (0, 16) | 1 (0, 2) |
| Outpatient Visits, median (IQR) | 52 (27, 88) | 160 (79, 286) | 63 (33, 116) |
| ECI, median (IQR) | 4 (2, 6) | 5 (3, 8) | 4 (2, 8) |
| CCI, median (IQR) | 1 (0, 2) | 2 (1, 3) | 2 (0, 3) |

<sup>a</sup> categorical variables reported as n (%)
<sup>b</sup> continuous variables reported as median (interquartile range)

Table 2 and Figures 1 and 2 report the meta-analysis results for the association between the MDD PGS and healthcare utilization and comorbidity burden. The average annual number of ED visits increased by 22.4% among those in the highest decile of MDD PRS compared to the lowest decile (RR=1.22, 95% CI: 1.17, 1.29). There was a 6.6% increase in average annual number of ED visits for each standard deviation (SD) increase in the MDD PGS (RR=1.07, 95% CI: 1.05, 1.08). Smaller effects were seen for IP visits, ECI and CCI (Table 1, Figures 1 and 2). The SCZ and BD PGS were not significantly associated with comorbidity burden, however there was a very modest though significant association with the number of ED visits for both BD (RR = 1.02, 95% CI: 1.01, 1.03) and SCZ (RR = 1.04, 95% CI: 1.02, 1.04) per each SD increase in PGS (Tables S5 and S6, Figures S1-S4).

**Figure 1.**
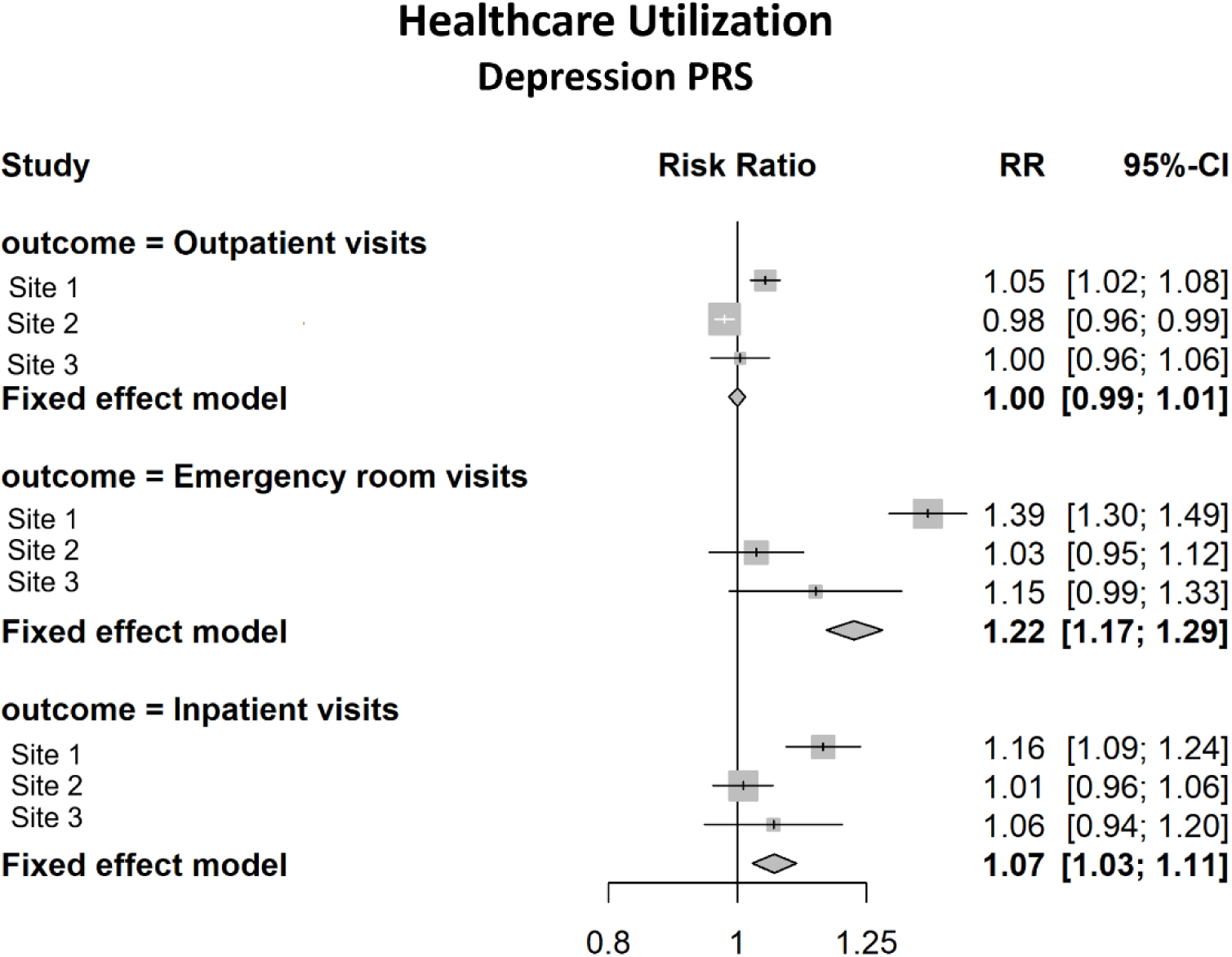
Meta-Analysis Results of the Association between 10^th^ and 1^st^ Decile of Major Depressive Disorder Polygenic Score and Healthcare Utilization Among Patients with no Depression Diagnosis

**Figure 2.**
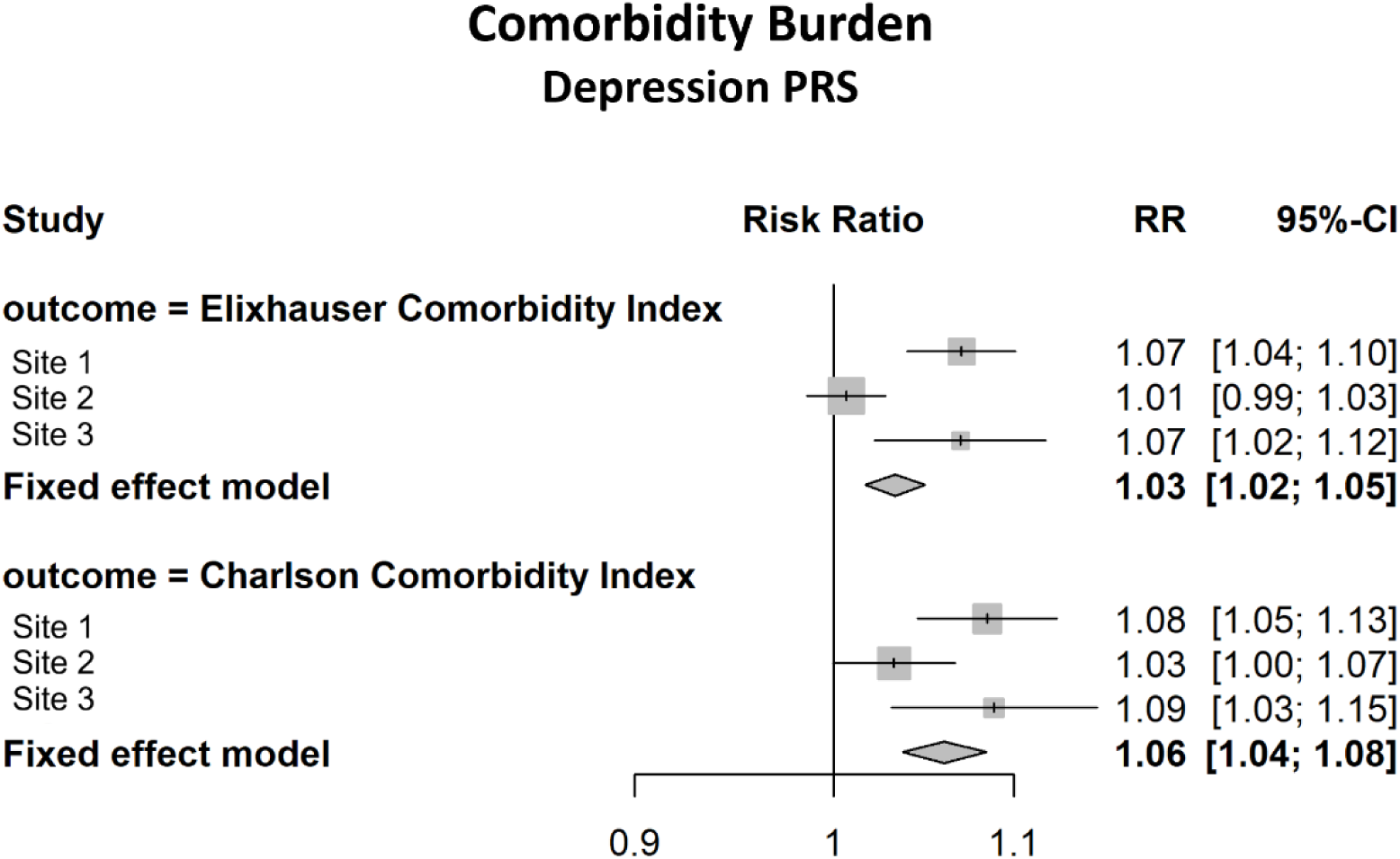
Meta-Analysis Results of the Association between 10^th^ and 1^st^ Decile of Major Depressive Disorder Polygenic Score and Comorbidity Burden Among Patients with no Depression Diagnosis

**Table 2:**
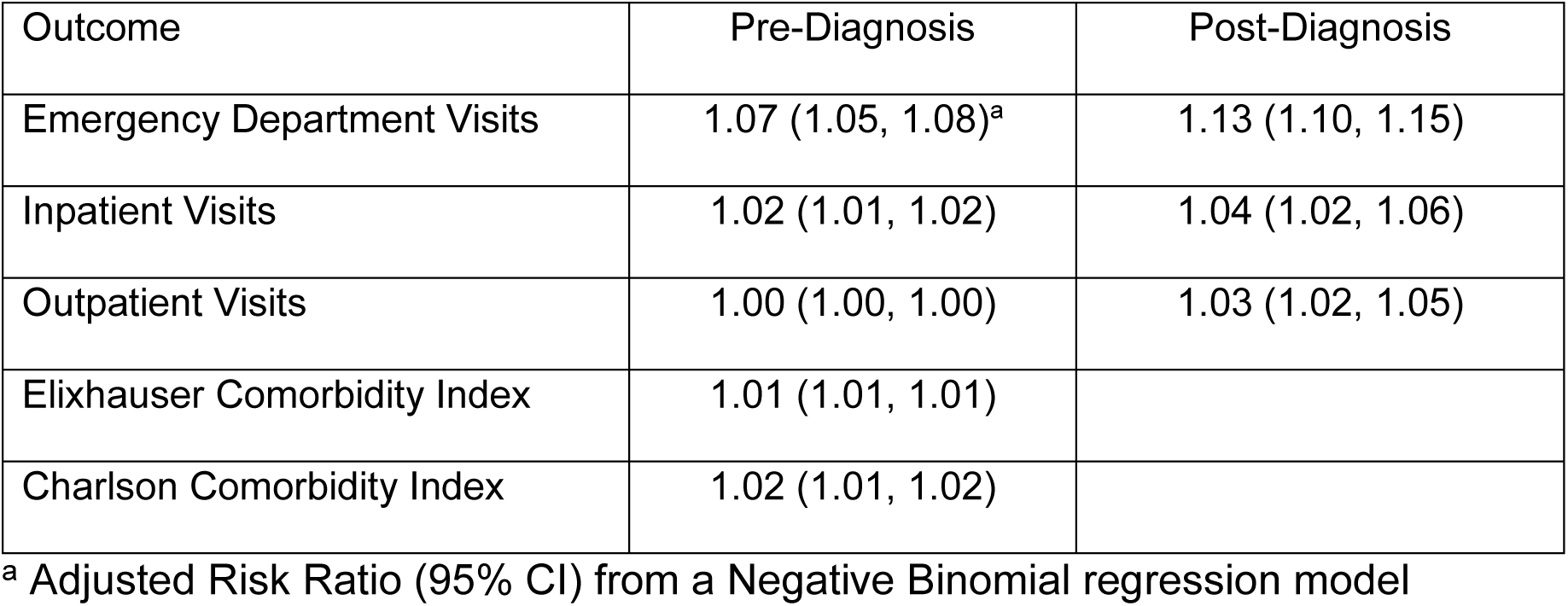
Meta-Analysis Results for Standardized Major Depressive Disorder Polygenic Score, Utilization and Comorbidity Burden.

| Outcome | Pre-Diagnosis | Post-Diagnosis |
| --- | --- | --- |
| Emergency Department Visits | 1.07 (1.05, 1.08)<sup>a</sup> | 1.13 (1.10, 1.15) |
| Inpatient Visits | 1.02 (1.01, 1.02) | 1.04 (1.02, 1.06) |
| Outpatient Visits | 1.00 (1.00, 1.00) | 1.03 (1.02, 1.05) |
| Elixhauser Comorbidity Index | 1.01 (1.01, 1.01) |  |
| Charlson Comorbidity Index | 1.02 (1.01, 1.02) |  |

<sup>a</sup> Adjusted Risk Ratio (95% CI) from a Negative Binomial regression model

Additionally, we examined the association between MDD PGS and healthcare utilization following a diagnosis of depression. In this subgroup analysis, there was a total of 53,595 participants meeting the definition of MDD across the three sites. Those in the highest PGS decile had a 56% increase in the average number of ED visits/year compared to the lowest decile (RR=1.55, 95% CI: 1.41, 1.72) (Figure 3). Similarly, there was a 13% increase per one SD increase in the standardized PGS (RR=1.13, 95% CI: 1.10, 1.15). Smaller but statistically significant associations were also found with OP and IP visits post-diagnosis. (Table 2).

**Figure 3.**
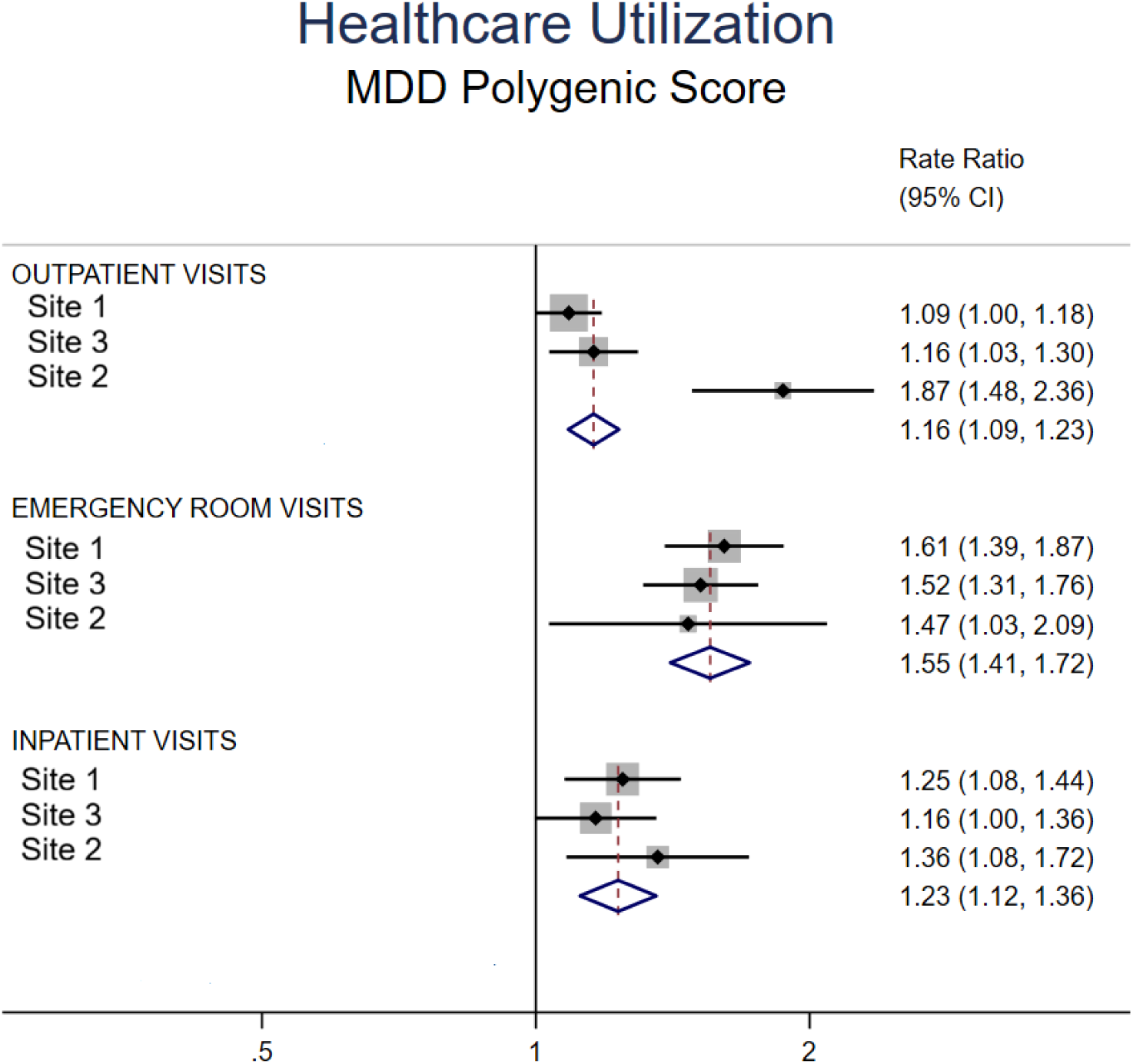
Meta-Analysis Results of the Association between 10^th^ and 1^st^ Decile of Major Depressive Disorder Polygenic Score and Healthcare Utilization after Depression Diagnosis

A sensitivity analysis was performed at Geisinger that excluded those participants that were coded as having MDD, SCZ, or BD. There were negligible changes in the results for SCZ and BD resulting in the same conclusion (data not shown). However, for associations with the MDD PGS, differences were found. The association comparing the highest to the lowest decile of the MDD PGS on utilization demonstrated no significant effect for OP, and the association with ED (RR=1.19, 95% CI: 1.13, 1.26) and IP (RR=1.03, 95% CI: 1.01, 1.05) were attenuated but remained significant.

## DISCUSSION

Psychiatric PGS have shown robust associations with psychiatric disorders^8,9,12^. However, less is known about their role as indices of healthcare utilization more generally. Here, we examined this issue by meta-analyzing data from EHR-linked biobanks across three large and geographically diverse healthcare systems in the PsycheMERGE network. Several notable findings emerged from these analyses. First, we found that, in the absence of a diagnosis of MDD, a PGS for MDD was significantly (though modestly) associated with ED and inpatient visits as well as two widely used indices of comorbidity burden (ECI and CCI). Following a diagnosis of depression, the MDD PGS was more strongly associated with ED visit frequency and was also associated with outpatient and inpatient visit frequency, suggesting that increased genetic liability to depression may be a marker of adverse prognosis among affected individuals. In contrast, both the SCZ and BD PGS showed attenuated and clinically insignificant associations with the healthcare outcomes.

The link between MDD PGS and both healthcare visits and chronic disease comorbidity may reflect pleiotropic effects of genomic contributions to depression. Prior reports have linked genetic risk for depression with a range of medical conditions, including cardiovascular, metabolic and immune-mediated disease^32-35^. Thus, individuals with greater genetic liability for depression might be expected to exhibit comorbidity with genetically correlated disorders for which they might also be more likely to seek care. Elevated healthcare resource utilization might also reflect a greater burden of subthreshold psychiatric and somatic symptoms that precede a formal depression diagnosis. We also find that utilization is even greater among those at higher levels of genetic risk after they receive a diagnosis of depression, suggesting that MDD PGS may also be a marker of worsened course among these patients. It is unclear why similar associations were not observed with PGS for SCZ or BD since these disorders have also been found to be genetically correlated with a range of psychiatric and medical syndromes^8^.

Our study has several strengths, including a sample size of over 110,000 individuals with both EHR and genomic data, with a median of 12 years of longitudinal EHR data.

The study also has limitations. We cannot accurately ascertain onset of mental health conditions; thus, some healthcare visits may have occurred after the development of the conditions of interest, but before documentation of diagnosis. To address this we performed a sensitivity analysis that excluded participants that were ever diagnosed with one of the conditions of interest. Significant associations persisted between MDD PRS and both ED and IP. Misclassification of diagnoses was possible if patients sought care outside of our health systems. Also, our effort to enhance specificity by requiring two or more depression diagnoses to define cases may have excluded true cases who received only one diagnosis. Finally, the effect sizes we observed were uniformly modest and there was some variability by site (i.e., smaller effects seen at VUMC).

Nevertheless, our study suggests that, in diverse real-world settings, polygenic contributions to depression may be biomarkers for healthcare resource utilization and comorbidity burden that could be lessened by preventative care, independent of diagnosed depression itself. We recognize the clinical utility of using PRS is limited given the modest effects found in our study. Future studies need to explore the combination of using PRS, along with other risk factors, to stratify patients at propensity for increased healthcare utilization.

## Data Availability statement

Protected Health Information restrictions apply to the availability of the clinical data here, which were used under IRB approval for use only in the current study. As a result, this dataset is not publicly available. Summary statistics are available by contacting the corresponding author.

## Author Contributions

Conceptualization: H.L.K., C.G.W., L.D., J.W.S; Data curation: H.L.K., D.R., R.K.L., M.R., H.L., Z.L., J.W.S.; Formal analysis: H.L.K., D.W., H.L.; Funding acquisition: J.W.S., L.D., C.F.C.; Writing-original draft: H.L.K., D.R., R.K.L, L.D., J.W.S.; Writing-review & editing: D.W., C.G.W., M.R., H.L., Z.L., Y.H., C.F.C.

## Ethics Declaration

The IRBs from Massachusetts General Brigham, Vanderbilt University Medical Center, and Geisinger have approved the study as non-human research due to de-identification of the data. All biobank participants gave informed consent for biobank research.

