## Supplemental Information for "Association Between Psychiatric Polygenic Scores, Healthcare Utilization and Chronic Disease Comorbidity Burden Among European Ancestry Individuals"

**Supplementary Information**

**Genotyping Information**

*Site 1*

In the data freeze used in the present study, Site 1 patient-participants were genotyped on either (a) the Illumina Human OmniExpress Exome BeadChip (OMNI; *N* ~ 59,000) or (b) the Illumina Global Screening Array (GSA; *N* ~ 74,500). The Regeneron Genetics Center imputed the genotypes to the reference panel of the Haplotype Reference Consortium^1^, by using the Minimac4 protocol on the Michigan Imputation Server^2^. As pre-imputation quality-control, SNPs were included based on (1) minor allele frequency (MAF) greater than 1%, (2) Hardy-Weinberg equilibrium (HWE) *P* value greater than 1×10^–15^, (3) variant missingness less than 1%. Samples were filtered on: (1) autosomal missingness greater than 2%; (2) excessive PC-adjusted heterozygosity; and (3) sex mismatch on the X chromosome (i.e., self-reported males with F_X_ less than 0.5 and self-reported females with F_X_ greater than 0.5). The imputed data was then subjected to an extensive quality-control procedure, described next.

To identify a genetically homogenous cluster of European-ancestry Site 1 samples, we generated principal components (PCs) with the 1000 Genomes the phase 3 version 5 reference data^3^. To select a set of high-quality SNPs, we first applied the following inclusion criteria on the 1000 Genomes data: (1) bi-allelic; (2) autosomal; (3) MAF greater than 1% among samples within each of the African (AFR), Admixed American (AMR), East Asian (EAS), European (EUR), and South Asian (SAS) population groups; and (4) not in any of the long-range LD regions reported in Price et al.^4^ and de Vlaming et al.^5^. The set of SNPs was further restricted to those that also met the following criteria within each of the imputed OMNI and GSA datasets: (1) bi-allelic and autosomal; (2) MAF greater than 1%; (3) imputation INFO score greater than 0.9; and (4) average maximum posterior call greater than 0.95.

At this stage, 2,828,598 SNPs remained in the 1000 Genomes data, which were used for relatedness pruning among reference samples belonging to the CEU, CHB, JPT, and YRI populations, similar to Bycroft et al.^6^. The samples were pruned using a KING kinship coefficient of 0.022^7^, after which 411 unrelated samples remained. The set of SNPs was further reduced to 1,695,781 SNPs that satisfied the following criteria within each of the relatedness-pruned groups of CEU, CHB, JPT, and YRI reference samples: (1) MAF greater than 5%; (2) HWE *P* value greater than 1×10^–6^; and (3) were not strand ambiguous (i.e., A/T or C/G SNPs). Finally, we applied LD-pruning with a sliding window of 1,000 SNPs, with an 80 SNP step-size, and an *r^2^* threshold of 0.1. The resulting set of 36,647 near-independent SNPs and the 411 unrelated 1000 Genomes samples were used for principal component analysis with PLINK1.9.

Lastly, we projected all unrelated 1000 Genomes samples (*N* = 2,243), as well as the Site 1 patient-participants, on the first four genetic PCs. This procedure identified a narrow cluster of Site 1 samples, which were selected based on being within the range on PCs 1–4 of 1000 Genomes samples belonging to the CEU, TSI, or GBR populations, and that self-reported to “White” and “Not Hispanic or Latino”. This procedure identified 124,467 narrowly defined European-ancestry samples. The principal components reported in this section was only used to identify a cluster of Europeans and the next section describes how we generated principal components that were included as control variables in the regression analyses.

To generate genetic PCs to be included as control variables in regression analyses, we first performed relatedness pruning of the OMNI and GSA datasets (KING coefficient of 0.022), using the same set of 1,695,781 SNPs as in the analysis above. A random draw of 6,421 unrelated narrowly defined European-ancestry samples was used for LD-pruning (same parameters as above), which identified 69,682 LD-pruned SNPs, which also had HWE > 5E-6 and geno < 0-95 a,pmg 24,679 unrelated OMNI samples, select for an initial round of PCA. Visual inspection of the PCs found that they now captured subtle population stratification that correlated with geography. Ancestral outliers that were further away than 5SD of the mean of PCs 1–10 was removed. Second round of PCA was performed to confirm that no ancestral outliers remained (final *N* = 122,370, of which 67,677 are unrelated), resulting in a set of 20 genetic PCs.

*Site 3*

Site 3 biobank (Site 3) participants were genotyped using the Illumina Multi-Ethnic Global array with hg19 coordinates. Site 3 methods followed the recommendations of Peterson et al. (2019)^8^. As pre-imputation quality-control, SNP-level QC filters removed variants with a call rate <98% and those that were duplicated across batches, monomorphic, not confidently mapped to a genomic location or associated with genotyping batch. For population assignment, sample-level QC filters removed individuals with a call <98%, MAF <95%, and strand ambiguous SNPs and long-range LD regions and pruned to <100K independent SNPs. To identify individuals with European ancestry, PCs of ancestry were calculated in the 1000 Genomes phase 3 reference panel and subsequently projected onto the Site 3 dataset, where a random forest classifier was used to assign ancestral group membership for individuals with a prediction probability >90%.

Within the European samples, sample-level QC filters identified unrelated individuals (p^ < 0.2) and removed individuals with a call <98%, excessive autosomal heterozygosity (±3 sd from the mean), or discrepant self-reported and genetically inferred sex. Within the unrelated European samples, PCs were calculated, and final SNP-level QC filters removed variants with call rate <98% and HWE <1e-10 and retained only autosomal SNPS, excluding indels and monomorphic SNPs. The Michigan Imputation Server was then used to impute missing genotypes with the Haplotype Reference Consortium dataset serving as the reference panel. Imputed genotype dosages were converted to hard-call format and subjected to further QC, where SNPs were removed if they exhibited poor imputation quality (INFO <0.8), low MAF (<1%), deviations from HWE (*P* < 1 × 10^−10^) or missingness (variant call rate <98%). Only unrelated individuals (p^ < 0.2) of EUR ancestry were included in the present study. These procedures yielded a final analytic sample of 24,826 individuals in the Site 3.

*Site 2*

The 98,473 individuals’ genetic data were genotyped by the Illumina Infinium expanded multi-ethnic genotyping array (MEGAEX), which contains 2,038,233 SNPs. SNP quality control steps include excluding SNPs with MAF < 0.005, Hardy-Weinberg equilibrium test P value ≤ 10^-10^ within each self-reported ancestry or call rate <95%. Individuals were removed if they had a mismatch between genetically inferred sex and self-reported sex, excess heterozygosity rate within each self-reported ancestry, missing rate ≥ 0.02, or potentially cross-contaminated samples (proportion IBD > 0.8). 94,369 samples and 887,250 high-quality autosomal SNPs remained.

Ancestry was determined with 1000 Genomes phase 3 (1000GP3) data. 1000 Genomes phase 3 (1000GP3) consists of 2,504 unrelated samples from 5 super populations African (AFR), Admixed American (AMR), East Asian (EAS), European (EUR), South Asian (SAS). 887,250 genotyped autosomal SNPs from the BioVU MEGAEX array was merged with 1000GP3 after removing C/G and A/T SNPs to avoid unresolvable strand mismatches in MEGA samples. Regions with known high LD^4^ (Chr 5 44–51.5 Mb, Chr 6 25–33.5 Mb, Chr 8 8-12Mb, Chr 11 45–57 Mb) were excluded and the common variants were then pruned (r2 < 0.05) using PLINK 1.9^9^ (–indep-pairwise 1000 50 0.05) to yield 71,339 SNPs in relative linkage equilibrium for ancestry analyses. Principal components (PCs) were generated using flashpca version 2.0. By using K nearest neighbors (KNN, k = 50) clustering, we inferred MEGA samples' ancestries. We treated 1000GP3 samples' PCs as the training set and MEGA samples' PCs as the test set. For each individual within the MEGA sample, we calculated its Euclidean distance to all reference individuals based on the 2 leading PCs and then identified the 50 nearest individuals. If at least 90% of the closest individuals were from the same super population, we inferred that the MEGA individuals belonged to that super population. Individuals not surpassing that threshold were considered admixed. Among the 94,369 MEGA individuals, 91,141 (96.6%) were assigned to a homogeneous super-population, with the following breakdown: AFR=14,176, AMR=1,152, EUR=74,612, EAS=762, SAS=439. The total set of individuals considered admixed was 3,228. A subset of individuals of EUR ancestries (MEGA-EUR) were selected for further analysis. All 94,369 samples were imputed on the Michigan Imputation Server v.1.2.4 using Eagle (V2.4.1) for phasing, Minimac4 for imputation and the Haplotype Reference Consortium (HRC) reference v1.1 panel in build GRCh37 as reference. Genotype probabilities were converted to hard-call genotypes using PLINK2 (hard-call ≥ 0.1). SNPs were filtered with imputation info score in any of the batches < 0.8 missing genotype rate > 0.02, or multi-allelic states (>2). Within EUR super populations, SNPs with MAF < 0.005 and Hardy-Weinberg equilibrium test P value < 1 × 10^-10^ were excluded.

**Polygenic risk scoring (PRS)**

Polygenic risk scores are a standard approach to collapsing aggregated risk from genome-wide association studies^10^. We tested for association of polygenic risk scores generated using the SNP weights from the most recent PGC summary statistics of relevant psychiatric traits including depression^11^, schizophrenia^12^ and bipolar disorder^13^ in Site 2 patients of European ancestry (MEGA-EUR). PRS analyses were performed using PRS-CS which places a continuous shrinkage prior on SNP effect sizes using a Bayesian regression framework^14^. The continuous shrinkage priors adapt the amount of shrinkage applied to each SNP to the strength of the associated GWAS signal based on the LD structure estimated from an external reference panel. Posterior SNP effects were generated in each cohort using PRS-CS and the 1000 Genomes European reference panel was used to estimate LD between SNPs. The PRS were calculated for each individual of the target cohort using Plink 1.9.

**List of ICD Codes for Major Depressive Disorder and Schizophrenia**

Table S1: List of Major Depressive Disorder ICD Codes

| ICD Version | Code | Description |
| --- | --- | --- |
| ICD9CM | 296.2 | Major depressive affective disorder, single episode, unspecified |
|  | 296.2 | Major depressive disorder, single episode |
|  | 296.22 | Major depressive affective disorder, single episode, moderate |
|  | 296.23 | Major depressive affective disorder, single episode, severe, without mention of psychotic behavior |
|  | 296.24 | Major depressive affective disorder, single episode, severe, specified as with psychotic behavior |
|  | 296.25 | Major depressive affective disorder, single episode, in partial or unspecified remission |
|  | 296.26 | Major depressive affective disorder, single episode, in full remission |
|  | 296.3 | Major depressive disorder, recurrent episode |
|  | 296.3 | Major depressive affective disorder, recurrent episode, unspecified |
|  | 296.32 | Major depressive affective disorder, recurrent episode, moderate |
|  | 296.33 | Major depressive affective disorder, recurrent episode, severe, without mention of psychotic behavior |
|  | 296.34 | Major depressive affective disorder, recurrent episode, severe, specified as with psychotic behavior |
|  | 296.35 | Major depressive affective disorder, recurrent episode, in partial or unspecified remission |
|  | 296.36 | Major depressive affective disorder, recurrent episode, in full remission |
| ICD10CM | F32 | Major depressive disorder, single episode |
|  | F32.0 | Major depressive disorder, single episode, mild |
|  | F32.1 | Major depressive disorder, single episode, moderate |
|  | F32.2 | Major depressive disorder, single episode, severe without psychotic features |
|  | F32.3 | Major depressive disorder, single episode, severe with psychotic features |
|  | F32.4 | Major depressive disorder, single episode, in partial remission |
|  | F32.5 | Major depressive disorder, single episode, in full remission |
|  | F32.8 | Other depressive episodes |
|  | F32.81 | Premenstrual dysphoric disorder |
|  | F32.89 | Other specified depressive episodes |
|  | F32.9 | Major depressive disorder, single episode, unspecified |
|  | F33 | Major depressive disorder, recurrent |
|  | F33.0 | Major depressive disorder, recurrent, mild |
|  | F33.1 | Major depressive disorder, recurrent, moderate |
|  | F33.2 | Major depressive disorder, recurrent severe without psychotic features |
|  | F33.3 | Major depressive disorder, recurrent, severe with psychotic symptoms |
|  | F33.4 | Major depressive disorder, recurrent, in remission |
|  | F33.40 | Major depressive disorder, recurrent, in remission, unspecified |
|  | F33.41 | Major depressive disorder, recurrent, in partial remission |
|  | F33.42 | Major depressive disorder, recurrent, in full remission |
|  | F33.8 | Other recurrent depressive disorders |
|  | F33.9 | Major depressive disorder, recurrent, unspecified |

Table S2: List of Schizophrenia ICD Codes

| ICD Version | Code | Description |
| --- | --- | --- |
| ICD9CM | 295 | Simple type schizophrenia |
|  | 295 | Simple type schizophrenia, unspecified |
|  | 295.01 | Simple type schizophrenia, subchronic |
|  | 295.02 | Simple type schizophrenia, chronic |
|  | 295.03 | Simple type schizophrenia, subchronic with acute exacerbation |
|  | 295.04 | Simple type schizophrenia, chronic with acute exacerbation |
|  | 295.05 | Simple type schizophrenia, in remission |
|  | 295.1 | Disorganized type schizophrenia, unspecified |
|  | 295.1 | Disorganized type schizophrenia |
|  | 295.11 | Disorganized type schizophrenia, subchronic |
|  | 295.12 | Disorganized type schizophrenia, chronic |
|  | 295.13 | Disorganized type schizophrenia, subchronic with acute exacerbation |
|  | 295.14 | Disorganized type schizophrenia, chronic with acute exacerbation |
|  | 295.15 | Disorganized type schizophrenia, in remission |
|  | 295.2 | Catatonic type schizophrenia, unspecified |
|  | 295.2 | Catatonic type schizophrenia |
|  | 295.21 | Catatonic type schizophrenia, subchronic |
|  | 295.22 | Catatonic type schizophrenia, chronic |
|  | 295.23 | Catatonic type schizophrenia, subchronic with acute exacerbation |
|  | 295.24 | Catatonic type schizophrenia, chronic with acute exacerbation |
|  | 295.25 | Catatonic type schizophrenia, in remission |
|  | 295.3 | Paranoid type schizophrenia |
|  | 295.3 | Paranoid type schizophrenia, unspecified |
|  | 295.31 | Paranoid type schizophrenia, subchronic |
|  | 295.32 | Paranoid type schizophrenia, chronic |
|  | 295.33 | Paranoid type schizophrenia, subchronic with acute exacerbation |
|  | 295.34 | Paranoid type schizophrenia, chronic with acute exacerbation |
|  | 295.35 | Paranoid type schizophrenia, in remission |
|  | 295.4 | Schizophreniform disorder, unspecified |
|  | 295.41 | Schizophreniform disorder, subchronic |
|  | 295.42 | Schizophreniform disorder, chronic |
|  | 295.43 | Schizophreniform disorder, subchronic with acute exacerbation |
|  | 295.44 | Schizophreniform disorder, chronic with acute exacerbation |
|  | 295.45 | Schizophreniform disorder, in remission |
|  | 295.5 | Latent schizophrenia, unspecified |
|  | 295.5 | Latent schizophrenia |
|  | 295.51 | Latent schizophrenia, subchronic |
|  | 295.52 | Latent schizophrenia, chronic |
|  | 295.53 | Latent schizophrenia, subchronic with acute exacerbation |
|  | 295.54 | Latent schizophrenia, chronic with acute exacerbation |
|  | 295.55 | Latent schizophrenia, in remission |
|  | 295.6 | Residual type schizophrenic disorders |
|  | 295.6 | Schizophrenic disorders, residual type, unspecified |
|  | 295.61 | Schizophrenic disorders, residual type, subchronic |
|  | 295.62 | Schizophrenic disorders, residual type, chronic |
|  | 295.63 | Schizophrenic disorders, residual type, subchronic with acute exacerbation |
|  | 295.64 | Schizophrenic disorders, residual type, chronic with acute exacerbation |
|  | 295.65 | Schizophrenic disorders, residual type, in remission |
|  | 295.7 | Schizoaffective disorder, unspecified |
|  | 295.7 | Schizoaffective disorder |
|  | 295.71 | Schizoaffective disorder, subchronic |
|  | 295.72 | Schizoaffective disorder, chronic |
|  | 295.73 | Schizoaffective disorder, subchronic with acute exacerbation |
|  | 295.74 | Schizoaffective disorder, chronic with acute exacerbation |
|  | 295.75 | Schizoaffective disorder, in remission |
|  | 295.8 | Other specified types of schizophrenia |
|  | 295.8 | Other specified types of schizophrenia, unspecified |
|  | 295.81 | Other specified types of schizophrenia, subchronic |
|  | 295.82 | Other specified types of schizophrenia, chronic |
|  | 295.83 | Other specified types of schizophrenia, subchronic with acute exacerbation |
|  | 295.84 | Other specified types of schizophrenia, chronic with acute exacerbation |
|  | 295.85 | Other specified types of schizophrenia, in remission |
|  | 295.9 | Unspecified schizophrenia |
|  | 295.9 | Unspecified schizophrenia, unspecified |
|  | 295.91 | Unspecified schizophrenia, subchronic |
|  | 295.92 | Unspecified schizophrenia, chronic |
|  | 295.93 | Unspecified schizophrenia, subchronic with acute exacerbation |
|  | 295.94 | Unspecified schizophrenia, chronic with acute exacerbation |
|  | 295.95 | Unspecified schizophrenia, in remission |
|  | V11.0 | Personal history of schizophrenia |
| ICD10CM | F20 | Schizophrenia |
|  | F20.0 | Paranoid schizophrenia |
|  | F20.1 | Disorganized schizophrenia |
|  | F20.2 | Catatonic schizophrenia |
|  | F20.3 | Undifferentiated schizophrenia |
|  | F20.5 | Residual schizophrenia |
|  | F20.8 | Other schizophrenia |
|  | F20.81 | Schizophreniform disorder |
|  | F20.89 | Other schizophrenia |
|  | F20.9 | Schizophrenia, unspecified |
|  | F25 | Schizoaffective disorders |
|  | F25.0 | Schizoaffective disorder, bipolar type |
|  | F25.1 | Schizoaffective disorder, depressive type |
|  | F25.8 | Other schizoaffective disorders |
|  | F25.9 | Schizoaffective disorder, unspecified |

**List of Medical Conditions included in the Comorbidity Indices**

A given condition was coded as positive if the corresponding ICD9/10 codes were present on at least two outpatient visits at least 30 days apart, or at least one emergency department visit, or at least one inpatient visit, or present on the Problem List, where available. Readers are referred to Quan *et al.^15^* for the list of ICD codes for each of the indices. Table S3 lists the individual medical conditions.

Table S3: List of Medical Conditions included in the Comorbidity Indices

| **Elixhauser Comorbidity Index** | **Charlson Comorbidity Index** |
| --- | --- |
| Congestive heart failure | Myocardial infarction |
| Cardiac arrhythmias | Congestive heart failure |
| Valvular disease | Peripheral vascular disease |
| Pulmonary circulation disorders | Cerebrovascular disease |
| Peripheral vascular disorders | Dementia |
| Hypertension, uncomplicated | Chronic pulmonary disease |
| Hypertension, complicated | Rheumatic disease |
| Paralysis | Peptic ulcer disease |
| Neurodegenerative disorders | Mild liver disease |
| Chronic pulmonary disease | Diabetes without chronic complication |
| Diabetes, uncomplicated | Diabetes with chronic compilation |
| Diabetes, complicated | Hemiplegia or paraplegia |
| Hypothyroidism | Renal disease |
| Renal failure | Any malignancy, except malignant neoplasm of skin |
| Liver disease | Moderate or severe liver disease |
| Peptic ulcer disease, no bleeding | Metastatic solid tumor |
| AIDS/HIV | AIDS/HIV |
| Lymphoma |  |
| Metastatic cancer |  |
| Solid tumor without metastasis |  |
| Rheumatoid arthritis/collagen vascular disease |  |
| Coagulopathy |  |
| Obesity |  |
| Weight loss |  |
| Fluid and electrolyte disorders |  |
| Blood loss anemia |  |
| Deficiency anemia |  |
| Alcohol abuse |  |
| Drug abuse |  |
| Psychosis |  |
| Depression |  |

**Supplementary Results for Bipolar and Schizophrenia Polygenic Scores**

Table S2 reports the descriptive statistics of the bipolar disorder (BD) and schizophrenia (SCZ) cohort stratified by site. A total of 118,882 biobank participants were included in the BD and SCZ analysis. The majority were female (54%-60%) and median duration in the EHR was at least 12 years in all sites.

Bipolar Polygenic Score

The BD polygenic score (PGS) was associated with an increase in emergency department (ED) visits for both the standardized and comparison of the highest to lowest decile. However, only the standardized BD PGS was significantly associated with ED visits. For each SD increase in the PGS, the number of ED visits increased by 2.0% (95% CI: 1.0%, 3.0%) (Table S5). There were no other significant associations (Figures S1 and S2).

Schizophrenia Polygenic Score

Similar results were observed for the SCZ PGS. For each SD increase in the PGS, the number of ED visits increased by 4.0% (95% CI: 2.0%, 4.0%) (Table S6). There were no other significant associations (Figures S3 and S4).

Table S4: Descriptive Statistics of Bipolar Disorder and Schizophrenia Analysis Cohort

| Variable | Site 1  (N=58,138) | Site 3  (N=24,826) | Site 2  (N=35,918) |
| --- | --- | --- | --- |
| Age, median (IQR)^a^ | 59.06 (16.86)^a^ | 61.68 (15.31) | 58.8 (16.75) |
| Female Sex, n (%)^b^ | 35,093 (60.4%) | 10,282 (53.8%) | 21,159 (58.9%) |
| EHR Duration (years), median (IQR) | 15 (10, 18)^b^ | 15 (10, 20) | 12 (8, 16) |
| Emergency Department Visits, median (IQR) | 1 (0, 3) | 0 (0, 3) | 0 (0, 1) |
| Inpatient Visits, median (IQR) | 1 (0, 2) | 7 (0, 25) | 1 (0, 2) |
| Outpatient Visits, median (IQR) | 57 (31, 96) | 197 (98, 355) | 64 (34, 119) |
| ECI, median (IQR) | 4 (2, 6) | 4 (2, 7) | 4 (2, 8) |
| CCI, median (IQR) | 1 (0, 3) | 1 (0, 3) | 1 (0, 3) |

^a^ categorial variables reported as n (%)

^b^ continuous variables reported as median (interquartile range)

Table S5: Meta-Analysis Results for Bipolar Polygenic Score,

Utilization and Comorbidity Burden

| Outcome | Standardized PGS | 10^th^ vs. 1^st^ Decile PGS |
| --- | --- | --- |
| Emergency Department Visits | 1.02 (1.01, 1.03)^a^ | 1.04 (0.99, 1.09) |
| Inpatient Visits | 1.00 (0.99, 1.01) | 0.99 (0.95, 1.02) |
| Outpatient Visits | 1.00 (1.00, 1.01) | 1.00 (0.99, 1.02) |
| Elixhauser Comorbidity Index | 1.00 (1.00, 1.00) | 1.01 (0.99, 1.02) |
| Charlson Comorbidity Index | 1.00 (0.99, 1.01) | 1.01 (0.98, 1.03) |

^a^ Meta-Analyzed Adjusted Risk Ratio (95% CI) per SD increase in PGS from a Negative Binomial regression model

Table S6: Meta-Analysis Results for Schizophrenia Polygenic Score,

Utilization and Comorbidity Burden

| Outcome | Standardized PGS | 10^th^ vs. 1^st^ Decile PGS |
| --- | --- | --- |
| Emergency Department Visits | 1.04 (1.02, 1.04)^a^ | 1.04 (0.99, 1.09) |
| Inpatient Visits | 1.01 (1.00, 1.02) | 1.01 (0.98, 1.05) |
| Outpatient Visits | 1.00 (1.00, 1.00) | 0.99 (0.98, 1.01) |
| Elixhauser Comorbidity Index | 1.00 (0.99, 1.00) | 0.98 (0.97, 1.00) |
| Charlson Comorbidity Index | 0.99 (0.99, 1.00) | 0.97 (0.95, 0.99) |

^a^ Meta-Analyzed Adjusted Risk Ratio (95% CI) per SD increase in PGS from a Negative Binomial regression model

Figure S1 – Meta-Analysis Results of the Association between 10^th^ and 1^st^ Decile of BD Polygenic Score and Comorbidity Burden
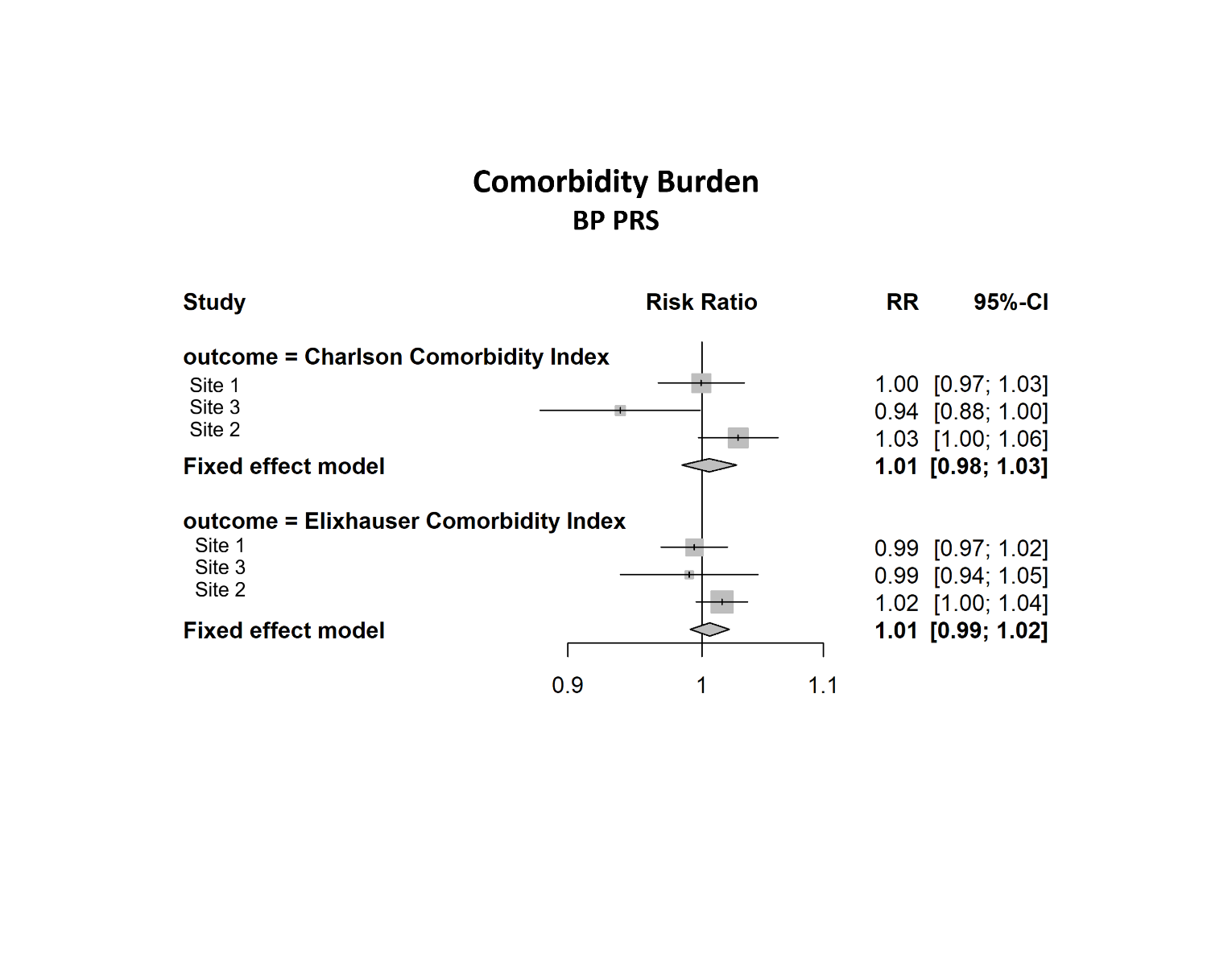

Figure S2 – Meta-Analysis Results of the Association between 10^th^ and 1^st^ Decile of BD Polygenic Score and Healthcare Utilization

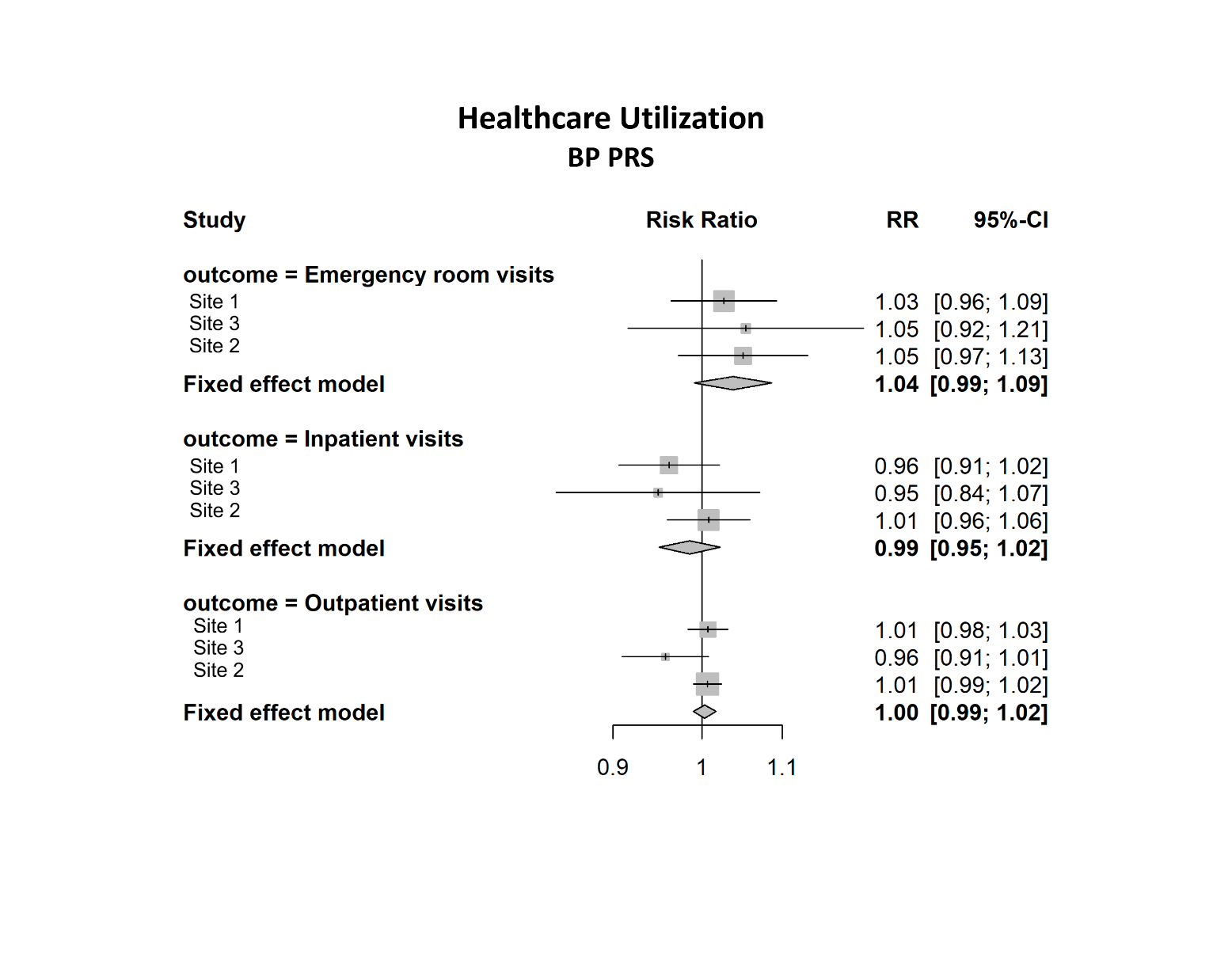

Figure S3 – Meta-Analysis Results of the Association between 10^th^ and 1^st^ Decile of SCZ Polygenic Score and Comorbidity Burden

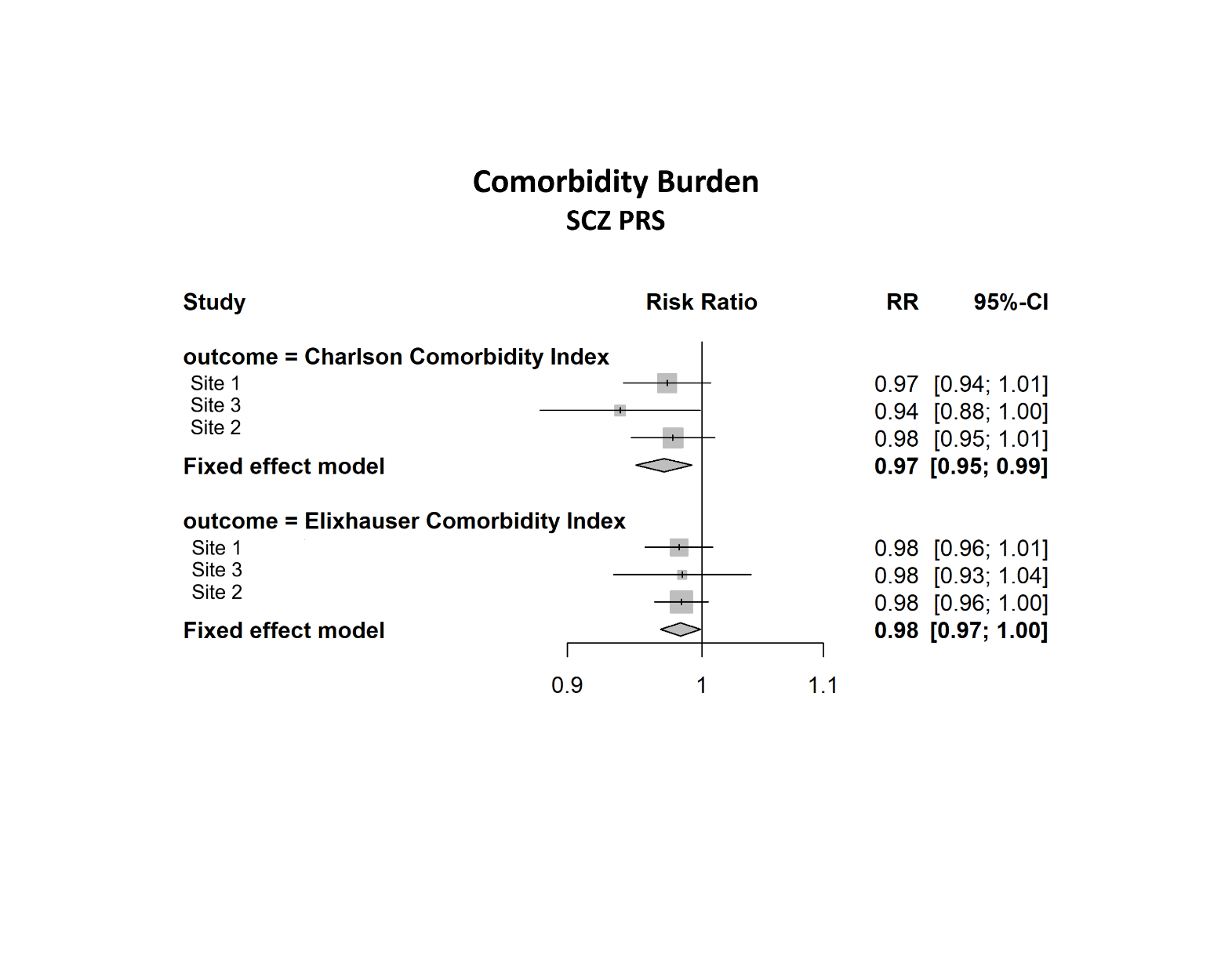

Figure S4 – Meta-Analysis Results of the Association between 10^th^ and 1^st^ Decile of SCZ Polygenic Score and Utilization

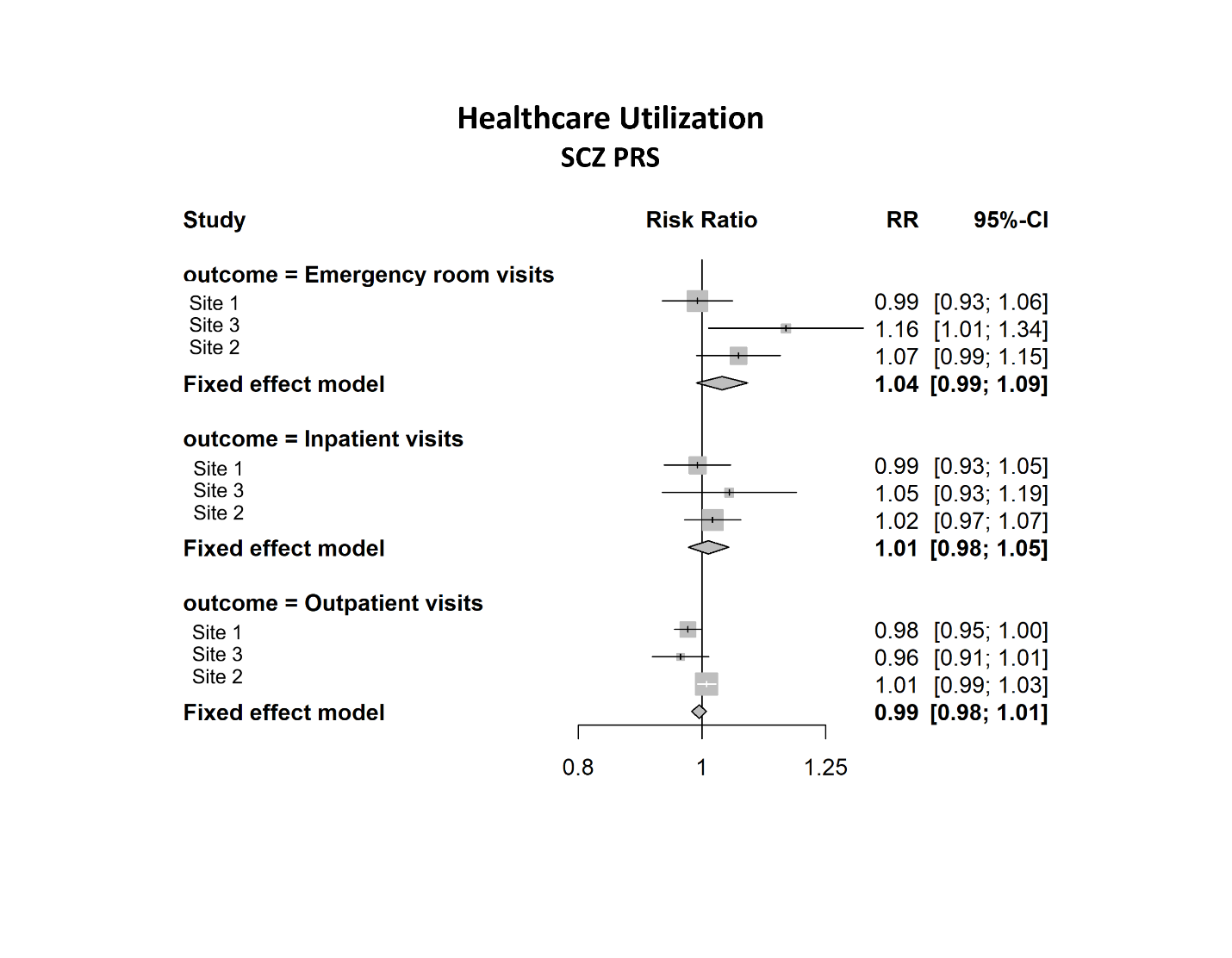
